# Isothermal Real-Time RT-RPA for Machupo Virus Detection: Field-Adaptable Sensitivity Comparable with Laboratory PCR

**DOI:** 10.1101/2025.08.22.25334221

**Authors:** Marina A. Kapitonova, Anna V. Shabalina, Igor S. Sukhikh, Artemiy A. Volkov, Vladimir G. Dedkov, Anna S. Dolgova

**Affiliations:** Laboratory of Pathogen Molecular Genetics, Saint Petersburg Pasteur Institute, St. Petersburg 197101, Russia; Martsinovsky Institute of Medical Parasitology, Tropical and Vector Borne Diseases, First Moscow State Medical University (Sechenov University), Moscow 119048, Russia

## Abstract

Isothermal nucleic acid amplification methods, such as recombinase polymerase amplification (RPA), are becoming increasingly vital as diagnostic platforms for neglected tropical diseases (NTDs) by enabling rapid and accurate detection in under-resourced regions. Wse developed a real-time RT-RPA assay for Machupo virus (MACV), the causative agent of Bolivian hemorrhagic fever, and directly compared it to a real-time RT-PCR assay targeting the same viral sequence fragment. The methods were evaluated across critical parameters: limit of detection (LOD), tolerance to single-nucleotide substitutions, multiplexing capability, and adaptability to multiple MACV genetic variants. The LOD was identical for both assays: 5×10^3^ copies/ml of armored RNA particles. They differed in terms of input RNA (copies/reaction): 100 (PCR) versus 20 (RPA). The real-time RT-RPA assay was further validated on a portable device, demonstrating its field-deployability for point-of-care applications.

**Author Summary:** We developed two new diagnostic tools to detect Machupo virus, which causes deadly Bolivian hemorrhagic fever in remote regions where lab resources are limited. Our tests, a lab-based RT-PCR and a portable rapid RT-RPA, both identify the virus with high sensitivity (detecting as few as 5,000 virus-mimicking particles per milliliter). The RT-RPA method works at a constant temperature, delivering results in 20–30 minutes on a handheld device, making it ideal for field clinics. While RT-PCR remains cost-effective ($1.15/test), RT-RPA ($5.95/test) offers unmatched speed and portability. Both tests perform well with viral genetic variability, ensuring reliable detection across strains. This work provides the first deployable molecular diagnostics tool for Machupo virus, filling a critical gap in neglected tropical disease outbreak response. Our study shows that rapid, field-ready diagnostics can match lab-based standards without sacrificing accuracy.

## 1. Introduction

Machupo virus (*Mammarenavirus machupoense*, MACV), a member of the *Arenaviridae* family, causes Bolivian hemorrhagic fever (BHF), a severe zoonotic disease endemic to rural Bolivia. First identified during a deadly 1963 outbreak with a 30% case fatality rate ^1^, MACV persists in nature through its rodent reservoir (*Calomys callosus*), with human infections occurring via aerosolized excreta or direct contact. BHF is classified as a neglected tropical disease (NTD). Like other hemorrhagic fevers caused by arenaviruses, it highlights the critical convergence of zoonotic spillover, limited healthcare access, and diagnostic gaps in resource-poor regions ^2^. Diagnosis of NTDs such as BHF presents systemic challenges: inadequate laboratory infrastructure; expensive equipment; and shortages of trained personnel in endemic areas.

Conventional MACV detection methods, serology (IgM/IgG ELISA) ^3^and lab-bound reverse-transcription PCR (RT-PCR) ^4^, cause critical delays in outbreak response in low-resource conditions. Therefore, the development and implementation of novel, rapid, and accurate point-of-care testing (POCT) methods becomes imperative ^5^. A key strategy involves isothermal nucleic acid amplification techniques, which enable rapid target detection using portable devices.

For instance, loop-mediated isothermal amplification (LAMP) has been successfully deployed for field identification of various bacterial ^6, 7^, parasitic ^8, 9^ and viral NTD pathogens. These include dengue ^10^, Zika ^11^, Lassa ^12^, Ebola ^13^, Marburg, Rift Valley fever, yellow fever, Congo Basin mpox, West African mpox, and variola viruses ^14^.

Another isothermal nucleic acid amplification method, rolling circle amplification (RCA), has been applied in diagnosis of the NTD black-grain eumycetoma by detecting six potential causative agents ^15^. This method has also been utilized for detecting type-specific bovine papillomavirus ^16^, H5N1 influenza virus ^17^, as well as the Ebola, dengue, and Zika viruses ^18^. Nucleic acid sequence-based amplification (NASBA) has demonstrated high efficacy in detecting diverse viral pathogens, including SARS-CoV-2 ^19^, rotavirus A ^20^, Zika virus ^21^, norovirus ^22^, and hepatitis B ^23^.

Among isothermal amplification methods, recombinase polymerase amplification (RPA) has gained significant traction in NTD diagnostics, demonstrating robust performance across diverse pathogens. It has been successfully deployed for parasitic targets ^24–28^, while equally enabling sensitive detection of high-consequence viruses, such as Crimean-Congo hemorrhagic fever virus (CCHFV) ^29^, severe fever with thrombocytopenia syndrome virus (SFTSV) ^30^, monkeypox virus ^31^, dengue virus ^32^, and Ebola virus ^33^.

However, despite the proven utility of isothermal methods for diverse NTD pathogen detection, no field-deployable molecular diagnostics exist for New World arenaviruses, particularly Machupo virus, which causes severe hemorrhagic fevers with pandemic potential. This critical gap leaves endemic regions without rapid tools for outbreak containment. To address this, we developed two methods: a real-time RT-RPA assay targeting a conserved region of the MACV RNA-dependent RNA polymerase gene; and a multiplex real-time RT-PCR assay for the same target. Here, we conducted a direct, head-to-head methodological comparison of isothermal RPA and conventional PCR methods. We analyzed sensitivity, cost, the influence of several single-nucleotide mutations on performance, and operational feasibility using armored RNA standards.

Furthermore, this comparative study addresses an important question in contemporary diagnostics: *Can rapid isothermal technologies like RPA realistically replace the gold-standard, PCR*? Despite its proven reliability, PCR can be less useful in time-sensitive outbreak scenarios where infrastructure is limited. By rigorously evaluating both assays under identical conditions, we provide empirical evidence for transitioning toward field-deployable platforms without compromising sensitivity. This work establishes the first portable, sequence-confirmed diagnostic for MACV while offering a transferable framework for arenavirus detection in resource-constrained settings.

## 2. Materials and methods

All primers for real-time PCR and RPA assays, along with fluorescent probes for RPA (Tables 1, S1), were synthesized by DNA-Synthesis (Moscow, Russia). Fluorescent probes for real-time PCR were synthesized by Genterra (Moscow, Russia). Probes for RPA contained a site-specific tetrahydrofuran (THF) residue, introduced as a custom modification by DNA-Synthesis (Moscow, Russia) using dSpacer CE Phosphoramidite (5’-O-dimethoxytrityl-1’,2’- dideoxyribose-3’-[(2-cyanoethyl)-(N,N-diisopropyl)]-phosphoramidite, Cambio, UK). All graphical visualizations, including kinetic plots, heatmaps, and standard curves were generated using MagicPlot 3.0.1 (MagicPlot Systems, Russia).

### 2.1. Positive Control Plasmid Construction

MACV target sequences (wild-type and variants) were synthesized *de novo* via PCR assembly using overlapping oligonucleotides, adapted from Dolgova and Stukolova ^34^. PCR was conducted using an outer pair of primers (M_1, M_4, 30 pmol) and inner primers (M_2, M_3, 1 pmol) with Phusion High-Fidelity DNA Polymerase (NEB, USA). The thermal cycling program was: 98°C for 30 sec; 27 cycles (98°C for 10 sec, 62°C for 30 sec, 72°C for 10 sec); and 72°C for 5 min. Products were purified using the DNA Clean & Concentrator Kit (Zymo Research, USA), A-tailed using Taq DNA Polymerase (Evrogen, Russia), and ligated into pGEM-T (Promega, USA). Plasmids were transformed into NEB Turbo *E. coli* (NEB, USA) and sequence verified.

Variant sequences (MACV_2, MACV_10, MACV_14) were synthesized identically using mutation-specific oligonucleotides (M_2_1-2, M_10_1-4, M_14_1-4). All inserts were subcloned into a linearized pET-MS2 vector, transformed, re-sequenced, and purified using the Plasmid Miniprep Kit (Zymo Research, USA). Linear DNA controls were generated by PCR amplification of plasmid inserts (2.3 kb target with flanking regions), purified using AMPure XP beads (Beckman Coulter, USA), and quantified via the NanoDrop One device (Thermo Fisher Scientific, USA).

### 2.2. Production and Purification of Armored RNA Particles (ARPs)

Recombinant plasmids were transformed into *E. coli* BL21(DE3). Protein expression was induced with 1 mM IPTG (neoFroxx, Germany) at 37℃ for 4 h. Cells were harvested by centrifugation, lysed (lysozyme treatment with freeze-thaw cycles), and digested with DNase I and RNase A (Zymo Research, USA). ARPs were purified from supernatants by CsCl (PanReac AppliChem, USA) density gradient centrifugation (200,000 ⅹ g, 22 h, 20℃). Primary fractions (3–4 per ARP variant) were screened using real-time RT-PCR with a mixed primer-probe system (Table 1). Optimal fractions were selected based on minimal Ct values: fraction 3 for MACV (Fig. S1); fraction 4 for MACV_2 (Fig. S2); and fraction 1 for MACV_10 (Fig. S3) and MACV_14 (Fig. S4).

**Table 1.**
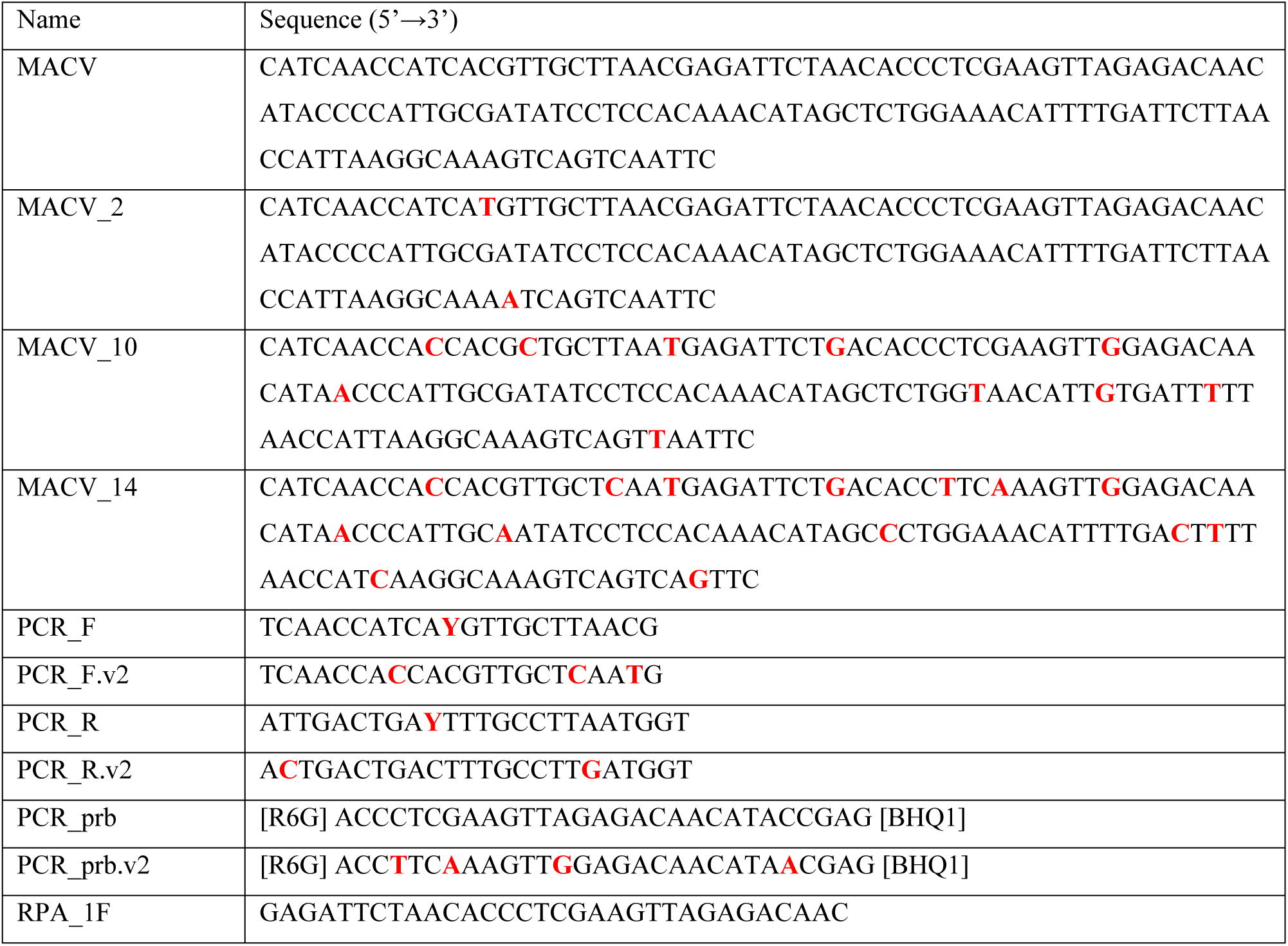
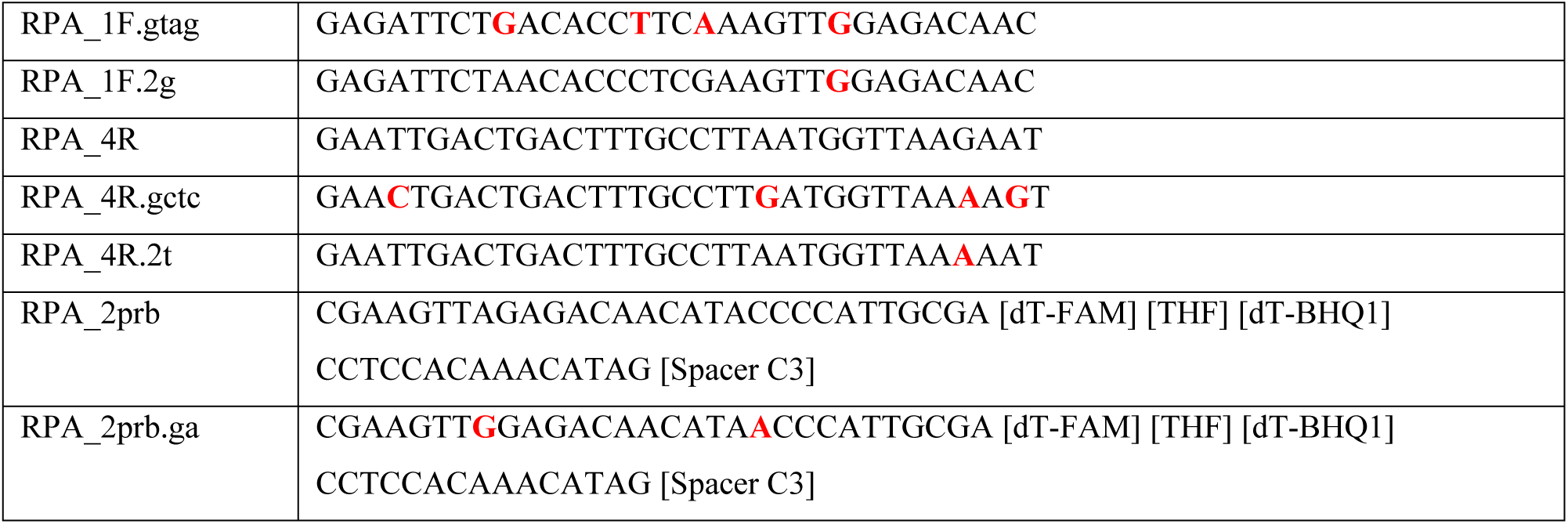
Target sequences and optimized primer/probe sequences (PCR/RPA). Nucleotide substitutions relative to MACV are highlighted in red.

### 2.3. RNA Purification and Droplet Digital PCR (ddPCR) Analysis

ARPs underwent seven 10-fold serial dilutions (10^-1^–10^-7^) in ultrapure water. Triplicate RNA extractions per dilution used the RIBO-prep Kit (AmpliSens, Russia). Purified RNA was immediately analyzed to prevent degradation. RNA quantification was performed using ddPCR (QX200™, Bio-Rad) with the One-Step RT-ddPCR Advanced Kit for Probes using flanking sequence-targeted primers/probes (SK_F/SK_R/SK_prb). Each reaction (20 μL) contained: 5 μl ddPCR Supermix; 2 μl reverse transcriptase; 1 μl 300 mM DTT; 1.8 μl each of 10 μM primers (SK_F, SK_R); 0.56 μl 10 μM SK_prb; 5.84 μl ultrapure water; and 2 μl RNA template. Reactions generated >10,000 accepted droplets, with thresholds manually set above no-template controls (NTC). Initial RNA concentrations were derived from averaged triplicates. Undiluted sample and extreme dilutions (10^-1^, 10^-6^, 10^-7^) were excluded as outliers. The log_10_-linear correlation between copy number and dilution factor is shown in Figure S5.

### 2.4. Real-Time RT-PCR

Optimized 25 μL reactions contained 12.5 μl of 2x RT-qPCR buffer (Biolabmix, Russia), 1 μl of 25x BioMaster mix (Biolabmix, Russia), 0.105 μl of each 100 μM primer (PCR_F, PCR_R, PCR_F.v2, PCR_R.v2), 0.075 μl of each 100 μM probe (PCR_prb, PCR_prb.v2), 0.93 µl DEPC-treated water (Biolabmix, Russia), and 10 μl RNA template. The aforementioned reaction mixture represents a primer/primer/probe concentration ratio of 7:7:5 per set. Alternative ratios (5:5:3 and 5:5:5) and annealing conditions (55/57/60℃ for 20/25/30 sec) were evaluated during optimization. The final thermocycling protocol on the CFX96 Touch (Bio-Rad, USA) was: 50℃ for 15 min; 95℃ for 5 min; and 40 cycles (95℃ for 10 sec, 60℃ for 30 sec) with HEX-channel detection.

### 2.5. Real-Time RT-RPA assay

**Primer Screening**. Initial primer combinations were evaluated using the TwistAmp® Basic kit (TwistDx™, UK) under standard conditions. Amplification products were directly loaded onto 2% agarose TBE gels stained with 1% ethidium bromide (PanReac AppliChem, USA). Electrophoresis ran at 80 V for 75 min (Mini-Sub Cell GT, Bio-Rad) alongside TriDye™ Ultra Low Range DNA Ladder (NEB, USA). Gels were imaged using a gelLITE Documentation System (Cleaver Scientific, UK).

**Optimized RT-exo RPA Protocol**. Reactions (50 μl) contained: 29.5 μl Primer Free Rehydration Buffer; 2.1 μl each of 10 μM forward/reverse primers; 0.6 μl of 10 μM fluorescent probe (THF-modified); 0.5 μl M-MuLV RT (200,000 U/ml, NEB, USA); 10.7 μl ultrapure water; 2.5 μl 280 mM magnesium acetate; and 2 μl RNA. After 40 seconds of vortexing, samples underwent isothermal amplification (40℃, 20 min) with minute-interval FAM-channel reads on a CFX96 Touch device (Bio-Rad, USA).

**Enzyme Variations**. For SuperScript™ IV RT (Invitrogen, 0.5 μl of 200 U/μl), RNase H (NEB, USA) was diluted in storage buffer to 0.5–2.5 U/μl. Storage buffer (pH 7.4) composition was: 50 mM KCl; 10 mM Tris-HCl; 0.1 mM EDTA; 1 mM DTT; 50% glycerol (PanReac AppliChem); and 200 μg/ml BSA (YACOO, China). Reactions used 2 μl RNA template and 1 μl RNase H, reducing water to 9.7 μl.

**Downscaled Screening & Portability**. Secondary screens used 25 μl reactions (all components halved). Field validation employed the Axxin T16-ISO device (Axxin, USA) with 30 minute amplification at 40°C.

## 3. Results

### 3.1. Target Sequence Selection

All available *M. machupoense* (MACV) complete L segment sequences were aligned using Mega v.11 software (Fig. S6). We chose a 136-nucleotide fragment of the RNA-dependent RNA polymerase gene as a target sequence. The reference sequence used was GenBank NC_005079.1 in the region from 1334 to 1469.

This sequence is unique for MACV, but exhibits variability across genetic variants. This deliberately selected non-conserved region was chosen to demonstrate a solution to a common challenge in diagnosing highly variable viruses. The selected sequence is conserved in the Carvallo strain (AY619642.1, JN794583.1, KM198593.1, AY216511.2, MT015969.1) and several isolates (KU978786.1, KU978789.1). However, it contains one or more single-nucleotide polymorphisms (SNPs) in the Chicava strain (AY624354.1, KU978785.1), the Mallele strain (AY619644.1, JN794585.1), and other isolates (KU978790.1, KU978805.1, KU978784.1, KU978787.1, KU978791.1, KU978788.1). Consequently, the reference sequence (MACV) and three variant sequences with 2, 10, and 14 nucleotide substitutions were selected as model targets. These were designated as MACV_2, MACV_10, and MACV_14 (representing KU978805.1, AY619644.1, and KU978787.1, respectively). Their alignment is shown in Figure 1 with binding sites for PCR and RPA primers/probes highlighted.

**Figure 1.**
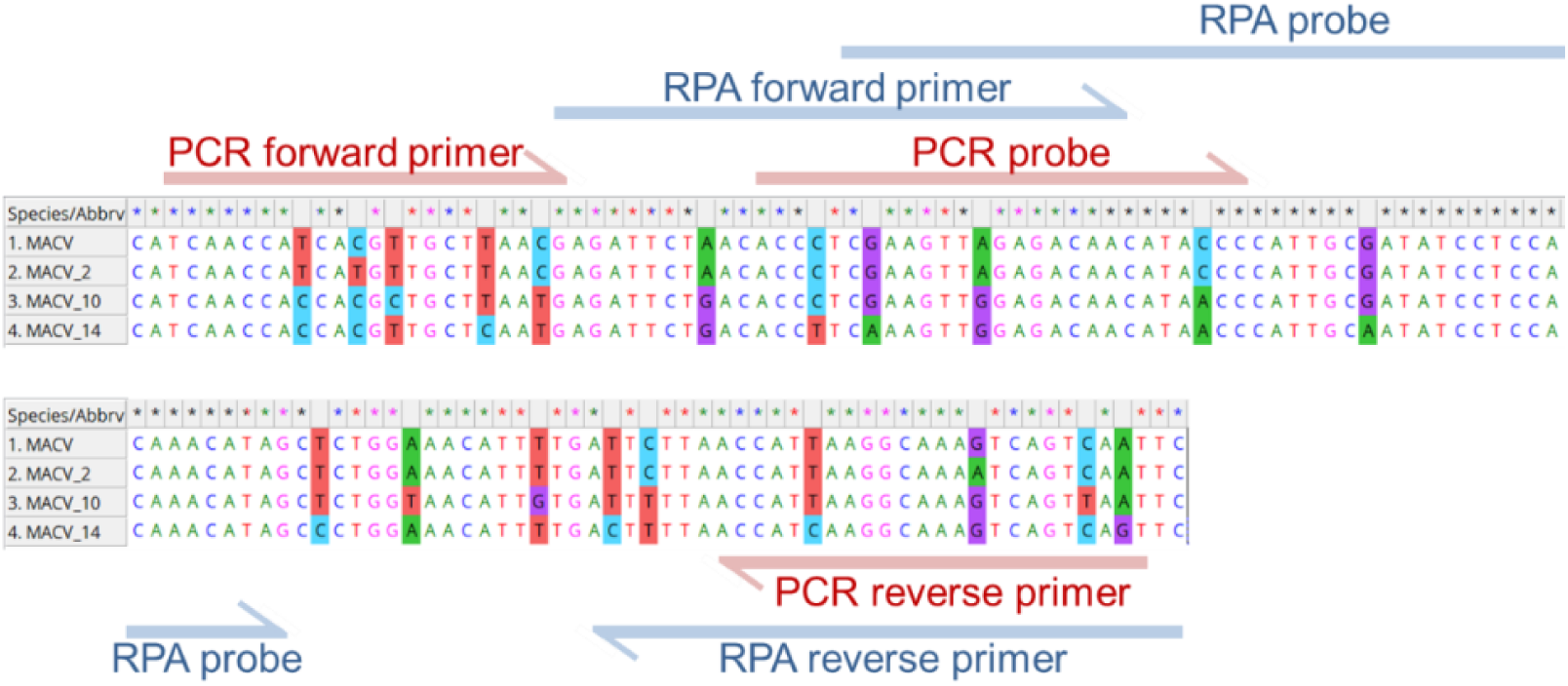
Sequence alignment of target variants. Nucleotide substitutions relative to MACV are highlighted. Binding sites for real-time RT-PCR and RT-RPA primers/probes are indicated.

### 3.2. Real-Time RT-PCR Assay

#### 3.2.1. Real-Time PCR Optimization

Preliminary optimization was carried out using a plasmid containing the MACV sequence. Primers PCR_F, PCR_R, and probe PCR_prb were designed to avoid dimerization and selected based on lowest Ct values. Optimization of primer and probe concentrations showed that the best ratio (forward/reverse/probe) was 7:7:5, yielding optimal performance (Table S2). Annealing temperature and duration optimization identified 60°C for 30 seconds as optimal (Table S3). The final protocol is detailed in the Materials and Methods section.

This primer-probe set was tested on plasmids containing the variant sequences (MACV_2, MACV_10, MACV_14). As expected, increasing numbers of substitutions correlated with higher Ct values, reflecting reduced reaction efficiency (Fig. S7). Amplification of MACV_14 was not detected.

To ensure detection of all variants, a second primer/probe set incorporating variant-matched substitutions (PCR_F.v2, PCR_R.v2, PCR_prb.v2) was introduced. Using a mixture of both primer sets, all control plasmids were successfully amplified (Fig. S8).

#### 3.2.2. Real-Time RT-PCR with ARPs

The optimized assay was tested on RNA extracted from MACV-containing ARPs, previously quantified by ddPCR. Seven ten-fold serial dilutions (5×10^7^ to 5×10^1^ copies/ml) of each ARP variant (MACV, MACV_2, MACV_10, MACV_14) were analyzed. The assay achieved a limit of detection (LOD) of 5×10^3^ copies/ml (100% replicate positivity) for all variants (Fig. 2). This equates to 100 target copies per reaction (10 copies/μl input RNA). As shown in Figure 2e, linear relationships between Ct value and log_10_(ARP concentration) were observed across the range 5×10^3^–5×10^7^ copies/ml (except 5×10^2^ for MACV_14).

**Figure 2.**
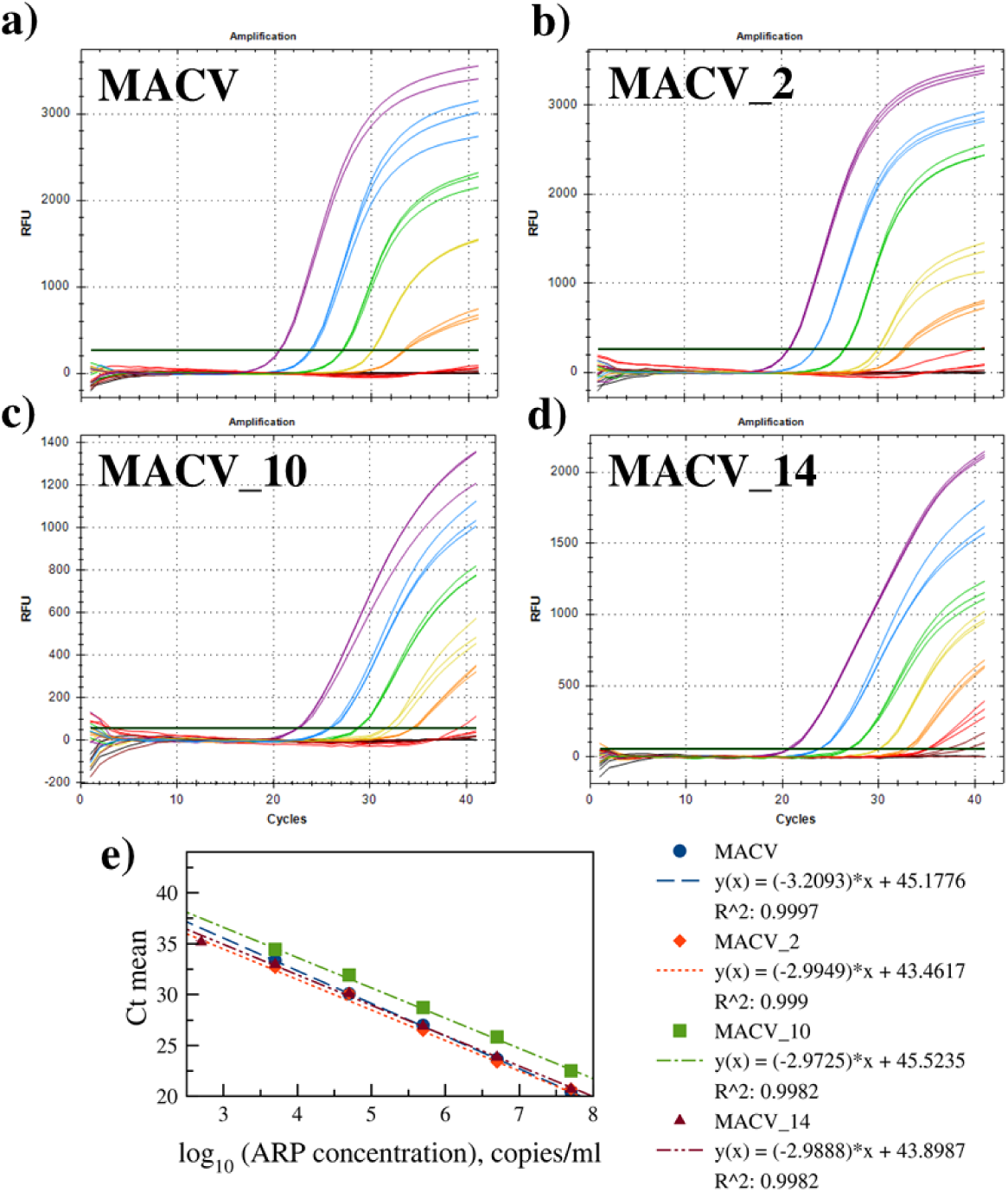
Real-time RT-PCR amplification plots using the mixed primer set on ARPs containing a) MACV, b) MACV_2, c) MACV_10, and d) MACV_14 target sequences. Serial 10-fold dilutions are shown (copies/ml): 5ⅹ10^1^ (brown), 5ⅹ10^2^ (red), 5ⅹ10^3^ (orange), 5ⅹ10^4^ (yellow), 5ⅹ10^5^ (green), 5ⅹ10^6^ (blue), 5ⅹ10^7^ (purple), and no template control (NTC, black). Panel e) shows standard curve with mean Ct values (± SD, n=3) versus log_10_(ARP concentration).

### 3.3. Real-Time RT-RPA Assay

#### 3.3.1. RPA Primer and Probe Selection

Recombinase polymerase amplification (RPA) efficiency critically depends on primer design. An initial screen of 2 forward (1F, 2F) and 3 reverse (4R, 5R, 6R) primers (6 combinations) assessed amplification by gel electrophoresis. The 1F/4R pair produced the highest-intensity amplicon band, suggesting superior efficiency (Fig. S9).

This pair was used for real-time RPA development. Two exo probes (1prb, 2prb), each containing a tetrahydrofuran (THF) residue and complementary to opposite strands, were designed. Both probes feature the motif [dT-FAM]-[THF]-[dT-BHQ1], enabling initial FAM fluorescence quenching by BHQ1 via proximity. Cleavage at THF by exonuclease separates the fluorophore-quencher pair, thereby releasing signal. Comparative testing identified probe 2prb as yielding higher fluorescence (Fig. 3a).

**Figure 3.**
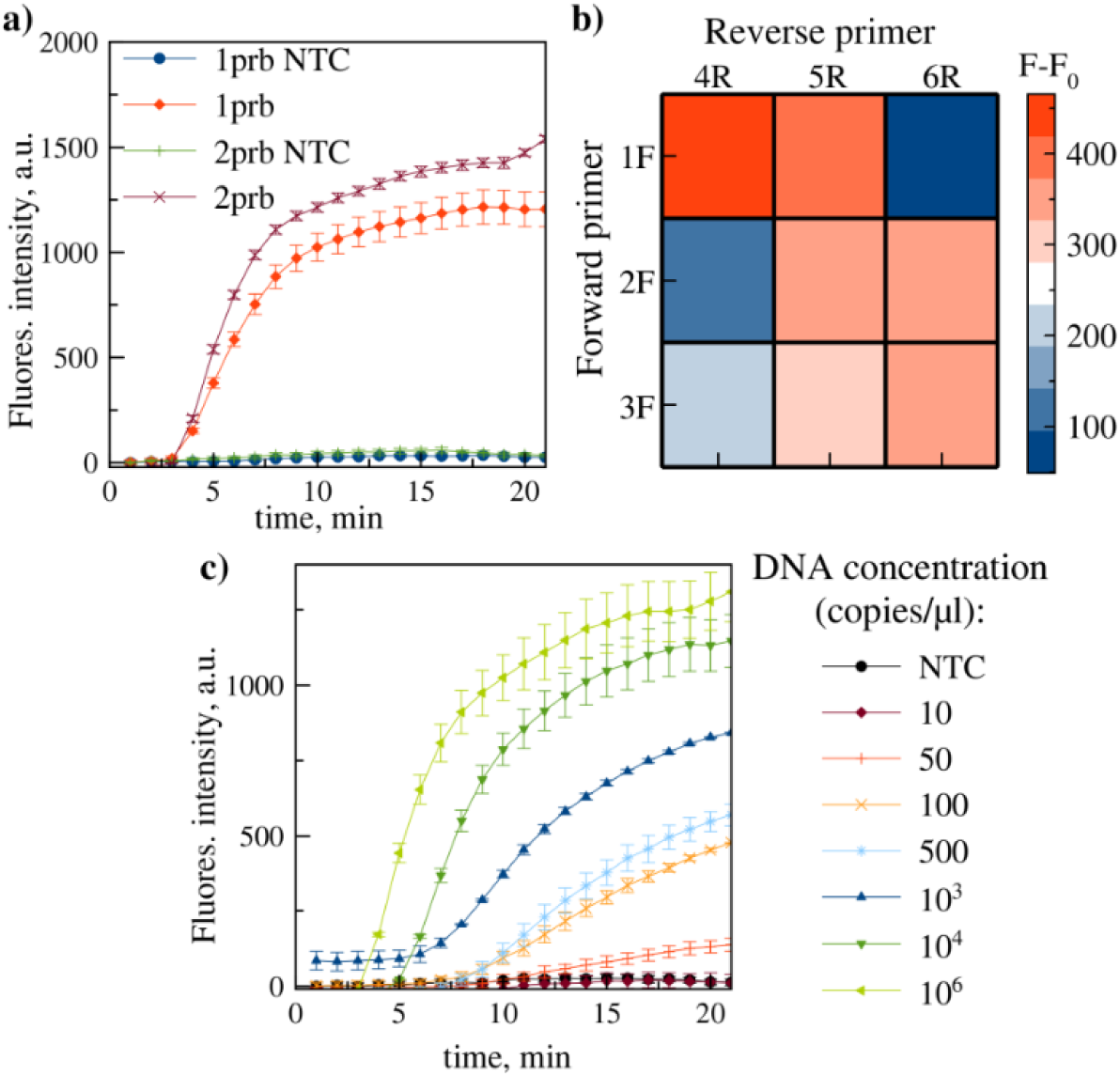
a) real-time RPA kinetics comparing probes 1prb and 2prb. b) heatmap of fluorescence intensity (F-F₀, background-subtracted) for primer pair screening (forward: 1F, 2F, 3F; reverse: 4R, 5R, 6R) with probe 2prb. c) real-time RPA kinetics using primer/probe set 1F/4R/2prb on serially diluted DNA template. Data represent mean ± SD (n=3 replicate reactions).

Primer/probe compatibility was further evaluated by screening the selected probe (2prb) with 3 forward (1F, 2F, 3F) and 3 reverse (4R, 5R, 6R) primers (9 combinations). Heatmap analysis confirmed 1F/4R as the optimal pair for 2prb (Fig. 3b). Sensitivity testing of the 1F/4R/2prb set on DNA template dilutions established a LOD of 10–50 copies/reaction (Fig. 3c).

#### 3.3.2. Testing Real-Time RT-RPA with ARPs

Assay performance with RNA was evaluated using MACV-containing ARPs (target sequence encapsulated within MS2 capsid, mimicking viral RNA structure). Reverse transcriptase (RT) was incorporated into the RPA mix. M-MuLV and its engineered mutant, SuperScript IV (offering higher processivity and thermostability), were compared. As SuperScript IV lacks intrinsic RNase H activity, exogenous RNase H was added. The optimal RNase H concentration was determined by testing 1 µl additions at 0, 0.5, 1.0, 2.5, and 5.0 U/µl concentrations. As shown in Figure 4a, the 0.5 U/µl concentration yielded maximal amplification efficiency and was selected for subsequent experiments.

**Figure 4.**
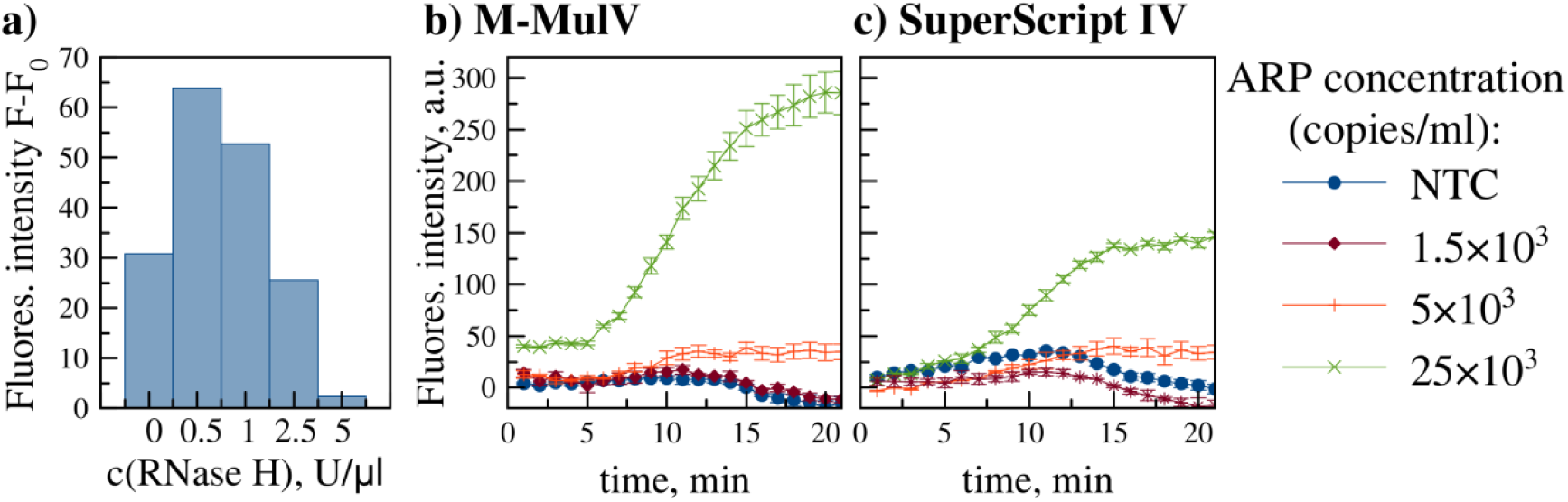
a) relative fluorescence (F-F_0_) at endpoint (20 min) for RT-RPA assays using SuperScript IV RT supplemented with varying concentrations of RNase H (0 - 5 U/µl). b) real-time RT-RPA amplification kinetics using optimized conditions (M-MuLV RT or SuperScript IV RT + 0.5 U/µl RNase H) and ARPs (1.5×10^3^ – 2.5×10^4^ copies/ml). Data represent mean ± SD (n=3).

Both RT systems achieved comparable LODs (5×10^3^ ARP copies/ml) as shown in Figure 4. However, M-MuLV generated significantly higher fluorescence at 2.5×10^4^ copies/ml and was selected as the preferred transcriptase.

#### 3.3.3. Optimization of RPA for Different Genetic Variants

The 1F/4R/2prb set showed reduced efficiency against variants: ∼40% lower fluorescence for MACV_2 and MACV_10; and no detection of MACV_14 (Fig. S10). To ensure robust detection across all genetic variants, two strategies were pursued. The first approach focused on rational primer redesign to incorporate compensatory point substitutions. Ideally, these minimize mismatches, while seeking a single “compromise” primer pair effective against all variants. We systematically screened 5 modified forward primers (1F variants) and 2 modified reverse primers (4R variants). Each contained from 1 to 3 nucleotide substitutions relative to the original sequences. Combinatorial screening results across the target templates are presented as a heatmap map (Fig. 5). Primer pair 1F.2g/4R.2t demonstrated the most balanced amplification performance and was selected as the optimal compromise candidate. However, testing this redesigned pair on ARP serial dilutions revealed a LOD of 5×10^4^ copies/ml (Fig. S11). This represents a 10-fold reduction in sensitivity compared to the original set (1F/4R/2prb) with the MACV target.

**Figure 5.**
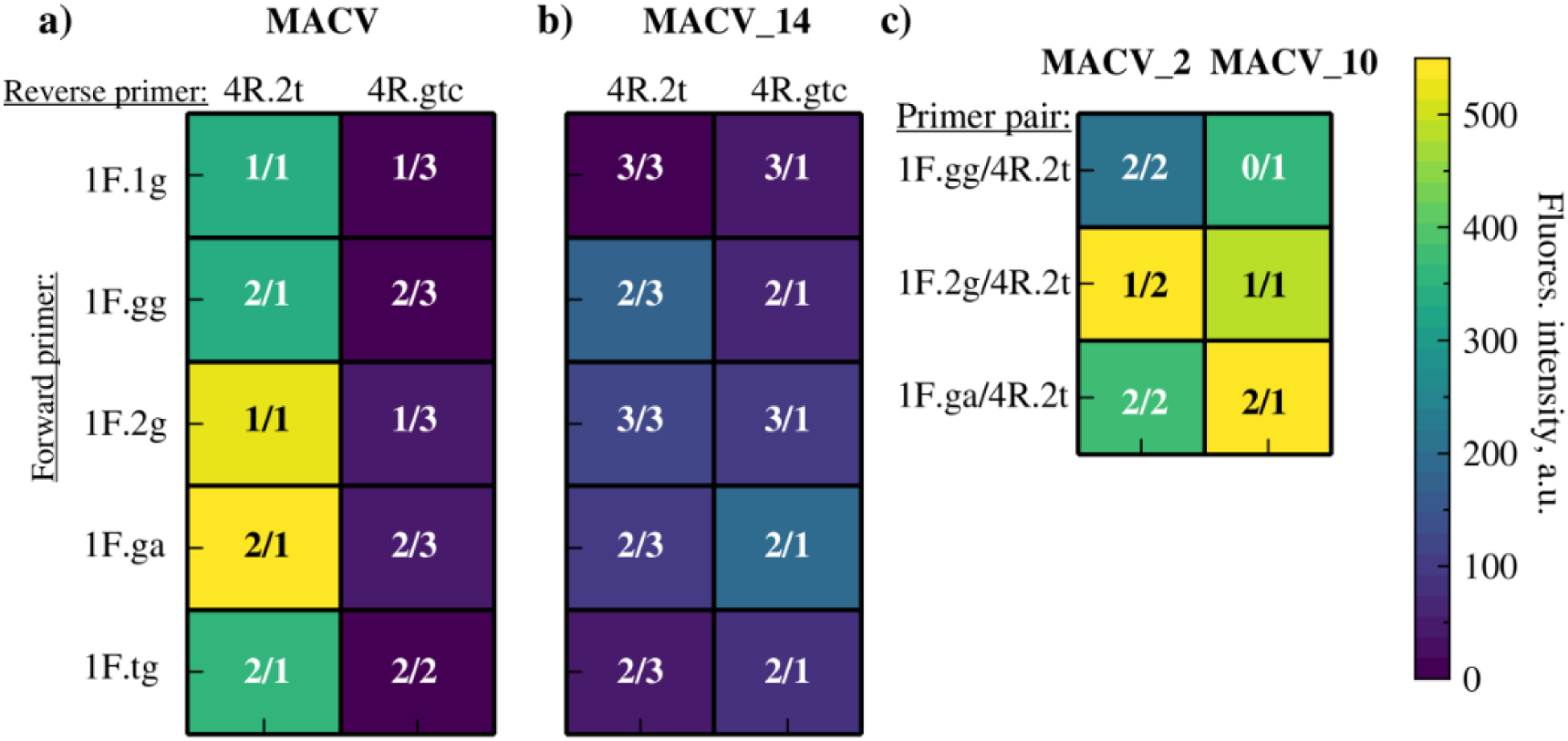
Heatmap of endpoint fluorescence intensity (F-F_0_) for redesigned primer pairs screened with probe 2prb across target variants. Primer variants: forward (1F.1g, 1F.gg, 1F.2g, 1F.ga, 1F.tg) and reverse (4R.2t, 4R.gtc). Numbers within cells (x/y) indicate substitutions in forward (x) and reverse (y) primers relative to original sequences. Panels: a) MACV, b) MACV_14, c) MACV_2 and MACV_10.

The second strategy, analogous to multiplex PCR approaches, involved adding a second, variant-specific primer-probe set designed to perfectly match MACV_14, the sequence with the highest substitution burden. This set consisted of primer ‘1F.gtag’, primer ‘4R.gctc’, and probe ‘2prb.ga’. Initial attempts to simply add this second set to the reaction mixture (resulting in a doubling of the total primer concentration) led to significant reaction inhibition (Fig. S12a). This issue was resolved by halving the concentrations of each individual primer (both original and new sets) within the combined mixture. This adjustment successfully overcame inhibition, while maintaining amplification capability (Fig. S12b).

Testing the optimized dual primer/probe mixture on ARP serial dilutions demonstrated an improved LOD of 5×10^3^ copies/ml (Fig. 6). This sensitivity matched the performance level previously achieved for real-time RT-RPA with the single primer set and was equivalent to the LOD obtained in the real-time RT-PCR assay.

**Figure 6.**
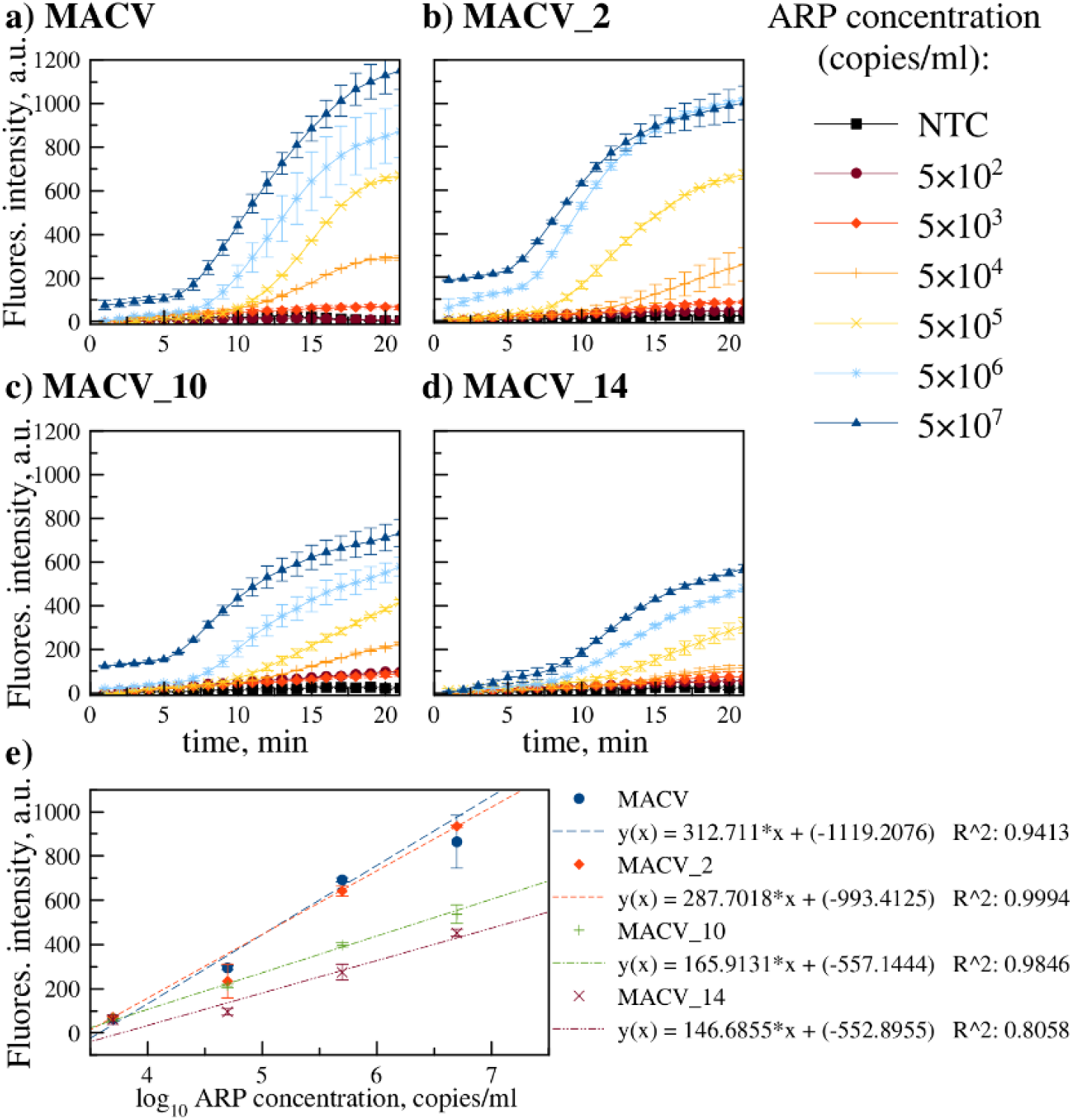
Real-time RT-RPA amplification kinetics with ARP serial dilutions containing: a) MACV, b) MACV_2, c) MACV_10, d) MACV_14. The optimized dual primer/probe set mixture was used (1F/4R/2prb + 1F.gtag/4R.gctc/2prb.ga, each at half concentration). LOD is indicated (5×10^3^ copies/ml). Panel e) linear dependence of fluorescence signal on log_10_(ARP concentration). Data represent mean ± SD (n=3).

Linear approximation in a specific concentration range (5×10^3^ – 5×10^6^ copies/ml) makes real-time RT-RPA (Fig. 6e) semi-quantitative, similar to real-time RT-PCR. However, it should be noted that with increasing concentration, the endpoint fluorescence value no longer fits this line due to reaction saturation. The linearity range in real-time PCR does not have this problem (Fig. 2e).

#### 3.3.4. Application of Real-Time RT-RPA with a Portable Device

Field-deployable diagnostics are essential for neglected tropical diseases like Bolivian hemorrhagic fever. We evaluated the optimized real-time RT-RPA assay on the battery-operated, isothermal Axxin T16-ISO fluorimeter using MACV ARPs. While the LOD remained 5×10^3^ copies/ml, extending the run time to 30 minutes (from 20) was necessary to maintain sensitivity (Fig. S13). Portable PCR systems, though available, were not assessed in this study.

## 4. Discussion

The persistent threat of neglected tropical diseases, such as Machupo virus infection, demands urgent innovation in diagnostic tools, particularly for pathogens with pandemic potential. As a zoonotic arenavirus transmitted from rodents to humans, Machupo exemplifies high-risk agents where genetic variability, driven by cross-species adaptation, can rapidly enhance transmissibility and virulence. With fatality rates exceeding 25%, and no licensed vaccines, early detection remains the primary barrier against outbreaks. Our study directly addresses this need through parallel development of real-time RT-PCR and RT-RPA assays. Both were rigorously compared using identical armored RNA targets and detection parameters to ensure equitable performance evaluation. The results of the comparison are briefly presented in Table 2.

**Table 2.** Comparative analysis of the real-time RT-PCR and RT-RPA assays.

| Parameter | Real-Time RT-PCR | Real-Time RT-RPA |
| --- | --- | --- |
| LOD (ARP copies/ml) | $5 \times 10^3$ | $5 \times 10^3$ (lab & portable) |
| LOD (RNA copies/reaction) | 100 | 20 |
| Time to result | 1 h 30 min | 20 min (lab), 30 min (portable) |
| Multiplex capability | yes (dual sets) | yes (dual sets) |
| Isothermal amplification | no | yes |
| Reaction cost (USD)† | 1.15 | 5.95 |
| Portable application | Palm PCR‡, not validated<br>(20 min, ~\$4.56) | Axxin T16-ISO, validated<br>(30 min) |
| Note: † excluding primers/probes; ‡ theoretical estimate using the Palm PCR™ Express One-step qRT-PCR Kit (Ahram Biosystems) and a pack of special tubes. The latter: type IIA Palm PCR™ real-time sample tubes. |  |  |

Both assays achieved clinically relevant sensitivity (LOD 5×10^3^ ARP copies/ml), while demonstrating robust detection across genetically diverse MACV strains. When translated to identical units (target input), RT-PCR detected 100 RNA copies/reaction, while RT-RPA showed enhanced efficiency at 20 RNA copies/reaction, reflecting its superior tolerance to low-input samples. The real-time RT-PCR protocol, optimized with dual primer/probe sets, provides a cost-effective solution (∼$1.15/reaction) for laboratories with existing infrastructure, delivering results in about 1 hour and 30 minutes. Critically, its multiplex design overcame challenges posed by highly variable targets (e.g., MACV_14 with 14 substitutions). In contrast, the RT-RPA assay, representing isothermal amplification, enabled field deployment, though at higher reagent costs (∼$5.95/reaction). When validated on the Axxin T16-ISO portable device, RT-RPA maintained its 5×10^3^ copies/ml sensitivity with only a 10 minute time extension (30 minutes total runtime).

While portable PCR systems like the Palm PCR™ could theoretically reduce RT-PCR runtime to 20 minutes at ∼$4.56/reaction, our study empirically confirms RT-RPA’s superior field adaptability. The Axxin-validated RT-RPA workflow eliminates thermal cycling dependencies and reduces power requirements; these are critical advantages in resource-limited settings.

Beyond conventional PCR and isothermal amplification (e.g., real-time RPA), emerging alternative nucleic acid detection platforms leverage nanotechnology, DNAzymes, and CRISPR-Cas systems. In our parallel study ^35^, we developed a ‘DNA endonuclease-targeted CRISPR trans reporter’ (DETECTR) assay targeting the identical MACV fragment using Cas12a. While promising, standalone DETECTR exhibited insufficient sensitivity (∼10^11^ copies/ml), necessitating RPA pre-amplification. The combined RT-RPA/DETECTR workflow achieved an LOD of 5×10^4^ ARP copies/ml with 60 min total runtime (30 min RPA + 30 min Cas12a detection). That represents an order-of-magnitude lower sensitivity compared to the real-time RT-RPA system reported here. Technically, RT-RPA/DETECTR proved more complex than the homogeneous real-time RT-RPA reaction insofar as the former requires spatial separation of reagents, namely: the RPA mix (at the bottom of the tube); and Cas12a reagents (at the cap).

Collectively, these studies provide the first direct comparison of PCR, RPA, and RPA-CRISPR platforms for arenavirus detection. Three key insights emerged. First, simplicity favors real-time RPA because single-tube, single-step protocols minimize operational errors in field settings. Second, speed-sensitivity trade-offs show that CRISPR-enhanced methods add complexity, without sensitivity gains, versus optimized RPA. Third, real-time RPA satisfies urgent frontline screening, while PCR remains for confirmatory testing. This methodological hierarchy aligns with the WHO’s recommendations for decentralized diagnostics: rapid frontline triage (RPA), followed by centralized verification (PCR).

## 5. Conclusion

This study successfully solves an important diagnostic problem in Machupo virus detection by developing and validating two complementary molecular approaches adapted to different healthcare settings. The real-time RT-PCR assay delivers laboratory-grade sensitivity (5×10^3^ ARP copies/ml, 100 RNA copies/reaction) at minimal cost, while utilizing multiplex primer-probe sets to overcome viral genetic diversity. Meanwhile, the real-time RT-RPA platform achieves equivalent analytical sensitivity in 20-30 minutes and can be applied in the field using portable devices, although at higher reagent cost.

RPA’s operational flexibility, combined with equivalent sensitivity to gold-standard PCR, positions it as a transformative tool for frontline Machupo surveillance. Future efforts should focus on reducing RT-RPA reagent costs and validating these assays against clinical samples in endemic regions. Accessible diagnostics are critical for timely control of zoonotic viral outbreaks, especially in dynamic settings featuring evolutionary changes.

## Data Availability

All data produced in the present work are contained in the manuscript and suppotring materials

## Author Contributions

Marina Kapitonova: Conceptualization, Formal analysis, Investigation, Methodology, Validation, Visualization, Writing – original draft. Anna Shabalina: Formal analysis, Investigation, Methodology, Validation, Writing – original draft. Igor Sukhikh: Formal analysis, Investigation, Methodology, Validation, Writing – review & editing. Artemiy Volkov: Conceptualization, Methodology, Writing – review & editing. Vladimir Dedkov: Supervision, Funding acquisition. Anna Dolgova: Supervision, Writing – review & editing.

## Conflict of Interest

The authors declare no conflict of interest.

## Data Availability

The data supporting this article have been included in the Supporting Information section.

## Supporting Information

**Table S1**. Complete list of oligonucleotide sequences with modifications.

**Figure S1**. Fraction selection for MACV armored RNA particles (ARPs) by real-time RT-PCR. Fluorescence kinetics for fractions 1 (blue), 2 (green), and 3 (purple). Optimal fraction (3) was selected based on lowest Ct value.

**Figure S2**. Fraction selection for MACV_2 armored RNA particles (ARPs) by real-time RT-PCR. Fluorescence kinetics for fractions 1 (blue), 2 (green), 3 (purple), and 4 (orange). Optimal fraction (4) was selected.

**Figure S3**. Fraction selection for MACV_10 armored RNA particles (ARPs) by real-time RT-PCR. Fluorescence kinetics for fractions 1 (blue), 2 (green), 3 (purple), and 4 (orange). Optimal fraction (1) was selected.

**Figure S4**. Fraction selection for MACV_14 armored RNA particles (ARPs) by real-time RT-PCR. Fluorescence kinetics for fractions 1 (blue), 2 (green), 3 (purple), and 4 (orange). Optimal fraction (1) was selected.

**Figure S5**. Linear fit analysis of ARP quantification by ddPCR. Log_10_-transformed concentrations of serial ARP dilutions (MACV) versus ddPCR-calculated RNA copies/ml. Outliers (undiluted, 10^-1^, 10^-2^, 10^-7^ dilutions) excluded.

**Figure S6.** Sequence alignment of Machupo virus L segment fragments. GenBank accession numbers: NC_005079.1 (reference), MT015969.1, KU978805.1, KU978791.1, KU978790.1, KU978789.1, KU978788.1, KU978787.1, KU978786.1, KU978785.1, KU978784.1, KM198593.1, JN794585.1, JN794583.1, AY624354.1, AY619644.1, AY619642.1, AY358021.2, AY216511.2.

**Table S2**. Primer/probe concentration optimization for real-time RT-PCR. Mean Ct values (Bio-Rad CFX Maestro) for MACV detection using varying concentrations of PCR_F, PCR_R, and PCR_prb. The optimal concentration ratio was 7:7:5.

**Table S3.** Annealing parameter optimization for real-time RT-PCR. Mean Ct values (Bio-Rad CFX Maestro) testing annealing temperatures (55-60℃) and durations (20-30 sec). Optimal conditions: 60℃ for 30 sec.

**Figure S7.** Real-time RT-PCR amplification for variant plasmids (initial primer set). Targets: MACV (blue), MACV_2 (green), MACV_10 (purple), MACV_14 (black), NTC (orange). Threshold: 0.05 (red). Note: MACV_14 was undetected.

**Figure S8.** Real-time RT-PCR with dual primer/probe sets. Successful detection of all variants: MACV (blue), MACV_2 (green), MACV_10 (purple), MACV_14 (black). Note: NTC (orange), auto-threshold (dark green).

**Figure S9**. RPA primer screening by gel electrophoresis. Lanes 1-6: amplicons from every primer pair (1F/4R, 1F/5R, 1F/6R; 2F/4R, 2F/5R, 2F/6R) with positive control. NTC 1-6: negative controls. Red highlight: RPA product from optimal 1F/4R pair (136 bp). Ladder: 10-700 bp.

**Figure S10**. Endpoint fluorescence of RT-RPA with MACV variants. Normalized signal for the 1F/4R/2prb set shows reduced detection of MACV_2/MACV_10 and failure to detect MACV_14.

**Figure S11**. RT-RPA sensitivity for redesigned primers (1F.2g/4R.2t/2prb). Serial ARP dilutions of (a) MACV, (b) MACV_2, (c) MACV_10, (d) MACV_14. LOD: 5×10⁴ copies/mL. Mean ± SD, n=3.

**Figure S12**. Dual primer/probe concentration optimization. Amplification kinetics with (a) full-concentration mix (inhibition) vs (b) half-concentration mix (optimal) for all variants.

**Figure S13.** Portable real-time RT-RPA validation on the Axxin T16-ISO device. Amplification kinetics for MACV ARPs using the optimized dual primer/probe mixture.

## Notes

### Competing Interest Statement

The authors have declared no competing interest.

### Funding Statement

This research was funded by the state program Ensuring chemical and biological safety of the Russian Federation.

